# Concurrent chemoradiotherapy combined with immunotherapy in locally advanced cervical cancer: an evaluation of efficacy

**DOI:** 10.64898/2026.11.25.26368911

**Authors:** Yanjun Xue, Yingqi Hao, Chaoli Shi, Xiaohui Sun, Kaili Li, Jinmeng Li

**Affiliations:** Gynecology, Anyang Cancer Hospital, Anyang, Henan Province, China

**Keywords:** LACC, programmed death-ligand 1 inhibitors, immunotherapy, concurrent chemoradiotherapy, biomarkers

## Abstract

**Objective:** This study examined whether adding sintilimab to standard treatment improves early tumor response and survival in locally advanced cervical cancer. We also determined how peripheral CD8^+^ T-cell counts and the tumor marker CA125 change from baseline to treatment completion, and whether these measures predict the benefit of combination therapy.

**Method:** This single-center retrospective cohort study included 126 patients with locally advanced cervical cancer (LACC; FIGO 2018 stage IB3–IVA) who received treatment at our institution between January 2023 and January 2025. Sixty-six received standard concurrent chemoradiotherapy (CCRT), and 60 received CCRT plus sintilimab (200 mg intravenously every 3 weeks throughout the treatment course). Peripheral blood samples were obtained at baseline and at treatment completion to quantify CD4^+^ and CD8^+^ T-cell proportions and serum CA125 levels. Short-term response was assessed by RECIST version 1.1. Progression-free survival (PFS) and overall survival (OS) were estimated by the Kaplan–Meier method and compared between groups with the log-rank test. Independent prognostic factors were identified using Cox proportional hazards regression, and likelihood ratio tests examined interactions between immune markers and treatment assignment.

**Results:** The objective response rate (ORR) was 81.67% with CCRT plus sintilimab versus 65.15% with CCRT alone (P = 0.045). Median progression-free survival (PFS) was 18.6 months in the combination group and 10.0 months in the CCRT group (log-rank P < 0.001). Median overall survival (OS) was not reached in the combination group but was 27.3 months in the CCRT group (log-rank P = 0.0027). After adjustment for age and BMI, combination therapy remained an independent favorable prognostic factor for both PFS (HR = 0.443, 95% CI: 0.290–0.678, P < 0.001) and OS (HR = 0.410, 95% CI: 0.230–0.731, P = 0.002).

Stratification by the median post-treatment peripheral CD8^+^ T-cell proportion (24.48%) gave a median PFS of 19.9 months in the high-CD8^+^ group versus 12.4 months in the low-CD8^+^ group (HR = 0.63, 95% CI: 0.42–0.94, P = 0.022). CD8^+^ T-cell stratification showed a significant interaction with treatment assignment for OS (P = 0.012). Using the median post-treatment CA125 level (31.25 IU/mL), median PFS was 11.9 months in the high-CA125 group versus 19.4 months in the low-CA125 group (HR = 1.89, 95% CI: 1.26–2.83, P = 0.0017); unlike CD8^+^ stratification, however, CA125 showed no significant interaction with treatment for either PFS (P = 0.959) or OS (P = 0.569).

**Conclusion:** In LACC, concurrent chemoradiotherapy combined with sintilimab improves objective response and is associated with longer PFS and OS. Patients with a higher post-treatment CD8^+^ T-cell proportion derive greater benefit from combination therapy, supporting its use as a stratification indicator of efficacy. CA125 mainly reflects tumor burden and overall prognosis rather than treatment benefit and is therefore unsuitable for selecting patients for immunotherapy.

## Introduction

Cervical cancer is the fourth most commonly diagnosed malignancy among women worldwide; GLOBOCAN 2022 estimates put the global burden at approximately 662,000 new cases and 348,000 deaths^1^. Under the FIGO 2018 staging system, stage IB3–IVA disease is defined as locally advanced cervical cancer (LACC)^2^, accounting for roughly 37%–60% of new diagnoses. Survival declines with advancing stage, alongside a higher incidence of lymph node metastasis and greater treatment resistance^3^. Cisplatin-based concurrent chemoradiotherapy (CCRT) is the standard curative treatment for LACC^4–7^. CCRT induces immunogenic cell death and releases damage-associated molecular patterns (DAMPs), thereby activating the host immune system^8,9^. Immune checkpoint inhibitors (ICIs) exploit this response by disrupting inhibitory checkpoint pathways, principally the programmed death-1 (PD-1)/programmed death-ligand 1 (PD-L1) axis and cytotoxic T-lymphocyte-associated protein 4 (CTLA-4); lifting this restraint allows T cells to mount a stronger antitumor response^10^. Radiation further remodels the tumor immune microenvironment by promoting antigen release and presentation and recruiting T cells into the tumor bed, which provides a mechanistic rationale for combining radiotherapy with PD-1/PD-L1 blockade^11^.

In advanced, recurrent, or metastatic cervical cancer, adding checkpoint blockade to chemotherapy has produced a survival benefit. In KEYNOTE-826, the 24-month overall survival rate among patients with PD-L1 CPS ≥ 1 was 53.0% in the pembrolizumab arm versus 41.7% in the control arm (HR = 0.60, 95% CI: 0.48–0.74)^12^. BEATcc reported a comparable advantage with atezolizumab, with median overall survival reaching 32.1 months against 22.8 months in the control group (HR = 0.68, 95% CI: 0.52–0.88)^13^. Despite these gains in the metastatic setting and ongoing refinements in local control, a substantial proportion of patients with LACC still relapse or develop distant metastasis after CCRT^14^. How to improve disease control and survival in this population without compromising radical chemoradiotherapy remains an open clinical question.

Peripheral blood lymphocyte subsets provide a noninvasive readout of systemic immune status, and their composition correlates with immune-cell infiltration in the tumor microenvironment and, ultimately, with clinical outcomes^15,16^. Most existing studies, however, have relied on a single baseline sample. Longitudinal paired measurements of immune and tumor markers before and after treatment, and their potential interactions with treatment regimens, remain largely unexplored.

In this study, we tracked changes in peripheral blood CD4^+^ and CD8^+^ T-cell proportions together with serum CA125 and SCC-Ag levels in patients with LACC receiving CCRT with or without sintilimab. We then assessed these markers as independent prognostic factors and potential effect modifiers of treatment response, with the aim of informing biomarker selection for future prospective trials.

## 1. Data and methods

### 1.1 Research Object and Ethical Approval

This single-institution retrospective cohort initially included 146 patients with locally advanced cervical cancer (LACC) treated at our hospital between January 2023 and January 2025. After predefined inclusion and exclusion criteria were applied, 20 patients were excluded for incomplete or lost follow-up records (n = 9), concurrent malignancies (n = 5), treatment interruption (n = 4), or distant metastasis confirmed before treatment initiation (n = 2). The remaining 126 patients formed the final study cohort. Eligibility required (1) age 18–70 years; (2) histologically confirmed cervical squamous cell carcinoma, adenocarcinoma, or adenosquamous carcinoma; (3) FIGO 2018 stage IB3–IVA; (4) treatment with CCRT alone or CCRT combined with sintilimab (200 mg intravenously every 3 weeks); and (5) no prior anticancer therapy, including surgery, chemotherapy, radiotherapy, immune checkpoint inhibitors, or systemic immunomodulatory agents.

Patients were excluded for (1) distant metastasis at initial diagnosis (FIGO stage IVB); (2) a concurrent or previous history of other malignancies; (3) active autoimmune disease requiring systemic immunosuppressive therapy; (4) systemic glucocorticoids at a prednisone-equivalent dose exceeding 10 mg/day for at least 2 consecutive weeks, or other immunosuppressive agents, within 4 weeks before treatment initiation; (5) active infection, including HIV, active hepatitis B or C, or active tuberculosis; or (6) severe cardiovascular disease or any other contraindication to CCRT.

This investigation was approved by the Medical Research Ethics Committee of Anyang Cancer Hospital (approval no. 2024KY17H01), and reporting followed the STROBE statement^17^. Each patient provided written informed consent before enrollment.

### 1.2 Study grouping and treatment plans

Treatment allocation. The 126 patients were divided into two groups. The control group (n = 66) received CCRT alone, consisting of pelvic external beam radiotherapy (EBRT) with concurrent weekly cisplatin (50 mg/m^2^ for six cycles). The combination group (n = 60) received the same CCRT regimen plus sintilimab (200 mg intravenously every 3 weeks), starting on day 1 of radiotherapy and continuing throughout the treatment course.

Both groups received the same radiotherapy protocol. EBRT was delivered with a 6-MV linear accelerator at 1.8–2.0 Gy per fraction, five fractions per week, to a total pelvic dose of 45–50 Gy over approximately 5 weeks. After EBRT, high-dose-rate intracavitary brachytherapy with iridium-192 was applied to the cervical primary tumor and any vaginal involvement, delivering 6 Gy per fraction to point A for a total of 30–35 Gy (5–6 fractions). The whole radiotherapy course was completed within approximately 8 weeks.

Pelvic MRI was performed at baseline and at treatment completion to measure the maximum tumor diameter. Peripheral venous blood was collected at the same time points for quantification of SCC-Ag, CA125, and T-lymphocyte subsets (CD4^+^ and CD8^+^).

### 1.3 Observe indicators

#### 1.3.1 General clinical data

Baseline characteristics were retrieved from medical records. Demographic and clinical variables included age, height, weight, BMI, histologic type, largest tumor diameter, and pelvic or para-aortic lymph node metastasis. Peripheral blood tests covered the serum markers SCC-Ag and CA125, and the T-cell subsets evaluated were CD4^+^ and CD8^+^ T cells.

#### 1.3.2 Clinical efficacy

Short-term efficacy was assessed according to RECIST 1.1^18^. Partial response (PR) was defined as an SLD reduction of at least 30% from baseline, maintained for more than 4 weeks, with no new lesions. Stable disease (SD) applied when lesions met neither the criteria for PR nor those for PD. Progressive disease (PD) was defined as an SLD increase of at least 20% together with an absolute increase of at least 5 mm, or the appearance of new lesions. The objective response rate (ORR) was calculated as CR + PR, and the disease control rate (DCR) as CR + PR + SD.

Sample collection and laboratory assays. Peripheral venous blood (3 mL per draw) was collected at two time points: within 3 days before treatment initiation and within 3 days after completion of CCRT. For T-lymphocyte subset analysis, samples were drawn into EDTA-anticoagulated tubes, gently inverted to mix, and processed by red blood cell lysis, antibody incubation, centrifugation, and PBS washing. CD4^+^ and CD8^+^ T-cell proportions were then quantified by flow cytometry. Serum SCC-Ag and CA125 levels were measured with a fully automated chemiluminescence immunoassay analyzer. The institutional upper reference limits were CA125 ≤ 35 IU/mL and SCC-Ag < 1.5 μg/L.

#### 1.3.3 Follow-up and survival endpoints are defined

Follow-up. Within one week of completing treatment, all patients were entered into the follow-up roster and individual records were opened. Follow-up combined outpatient visits with telephone contact. For the first two years after treatment, patients were assessed every 3 months and thereafter every 6 months. At each visit, survival status, local recurrence, and distant metastasis were recorded.

Response assessment. Imaging was performed 8–12 weeks after treatment, and objective tumor response was evaluated according to RECIST 1.1.

At the same time, pelvic color Doppler ultrasound, chest CT, and a gynecological physical examination were performed, along with ECG and liver and kidney function tests. PFS was defined as the time from treatment initiation to disease progression (per RECIST 1.1) or death from any cause; patients without progression were censored at the last follow-up. OS was defined as the time from treatment initiation to death from any cause; surviving patients were censored at the last follow-up. All 126 patients in the final analysis completed the scheduled follow-up, and their records were available in full. Patients with incomplete follow-up information or those lost to follow-up had been excluded beforehand under the exclusion criteria. This study focused on efficacy, survival, and changes in peripheral blood markers and did not include a systematic statistical analysis of adverse reactions. Figure 1 outlines patient flow from screening and group assignment through follow-up.

**Figure 1.**
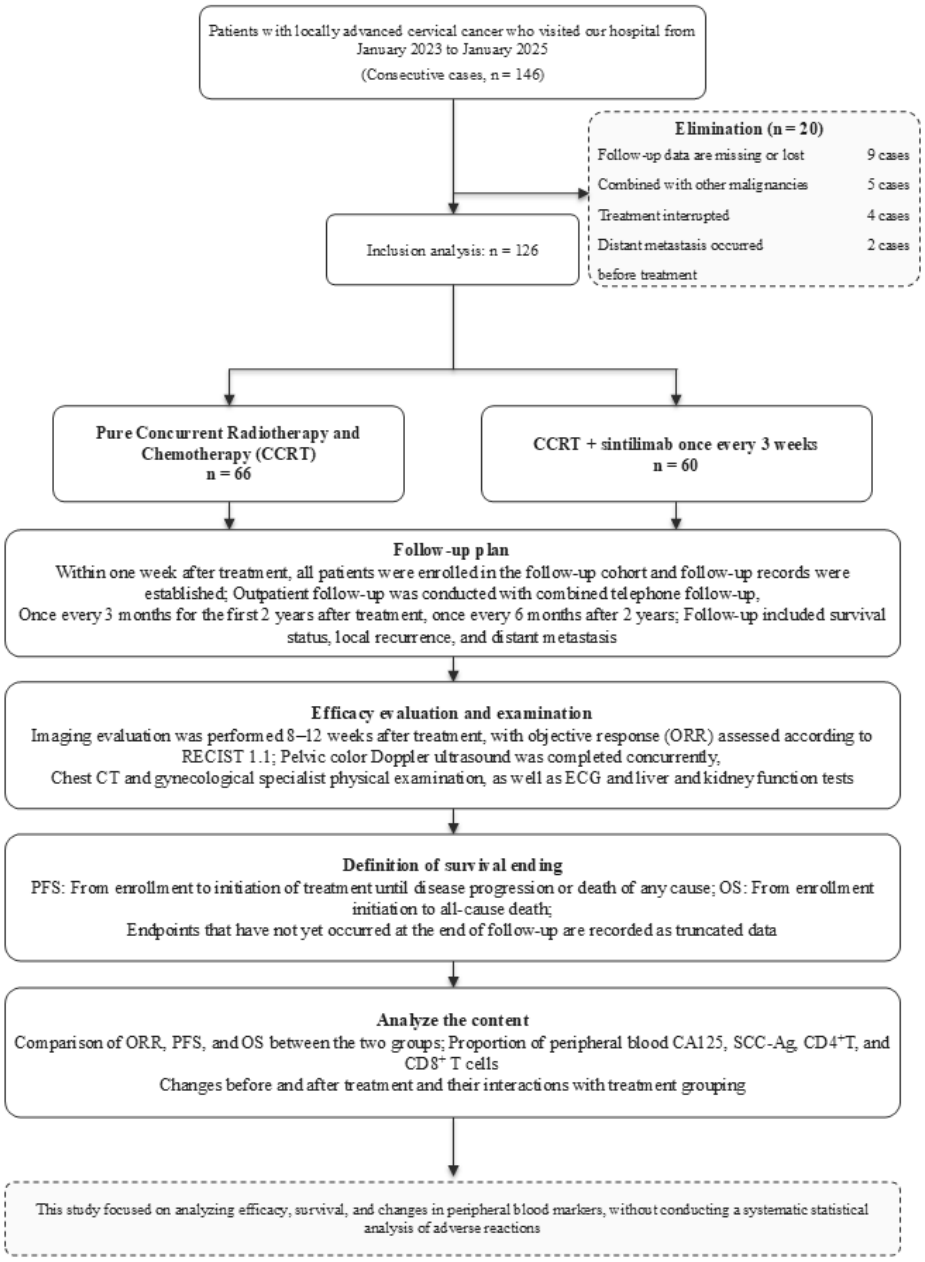
Flowchart of subject screening, grouping and follow-up

### 1.4 Statistical methods

Statistical analyses were performed in SPSS 26.0 and R (version 4.x). Continuous variables are reported as mean ± standard deviation. Between-group comparisons used the independent-samples t-test, with Welch’s correction applied when variances were unequal. Categorical variables are summarized as counts (percentages) and compared with the χ^2^ test; where the minimum expected frequency was below 5, Fisher’s exact test was used instead, and no continuity correction was applied to the 2×2 tables. Two-sided P < 0.05 was considered statistically significant.

PFS and OS were estimated by the Kaplan–Meier method, and curves were compared between groups with the log-rank test. The 24-month survival rate was reported for each endpoint. Median post-treatment CA125 and CD8^+^ T-cell values were used as cutoffs for stratified analyses. Cox proportional hazards regression was used for univariate and multivariate analyses; continuous variables were entered at a one-standard-deviation scale (per SD) so that differences in units would not affect interpretation of the hazard ratio (HR). Treatment group, together with age, BMI, post-treatment CA125, and post-treatment CD8^+^ T cells, entered a multivariable Cox model, from which adjusted HRs and 95% CIs were obtained. Discrimination was summarized with Harrell’s C-index, and Schoenfeld residuals were used to check the proportional hazards assumption. Interactions between stratification factors and treatment were evaluated with the likelihood ratio test by comparing nested models that differed only in the inclusion of the product term. A 3-month landmark analysis was then performed: cases progressing or dying within the first 3 months were removed before survival was recomputed, preventing early events from distorting subsequent risk estimates. Finally, the restricted mean survival time (RMST) difference was calculated over a 36-month horizon (τ = 36), a measure that does not rely on proportional hazards.

## 2. Results

### 2.1 General data comparison between the two groups of patients

A total of 126 patients with cervical cancer entered the analysis: 66 in the control arm and 60 in the immunotherapy arm. Baseline indicators including height, weight, BMI, CA125, SCC, CD4^+^ T lymphocytes, CD8^+^ T lymphocytes, and the CD4^+^/CD8^+^ ratio did not differ significantly between the groups (P > 0.05), indicating comparable baseline characteristics.

Mean age was 61.53 ± 3.47 years in the control group and 63.08 ± 3.14 years in the combination group, a significant difference (t = −2.641, P = 0.009). With a cutoff of 60 years, the combination group contained a higher proportion of patients older than 60 years (80.0% vs. 62.1%, χ^2^ = 4.843, P = 0.028). Age was therefore entered as a covariate in subsequent comparative analyses.

### 2.2 Differences in tumor markers and T lymphocyte subsets in peripheral blood were compared between the two groups before and after treatment

Table 2 compares pre- and post-treatment changes in the measured indicators between the two groups. Because age was unevenly distributed at baseline, age and the pre-treatment value were entered as covariates in an analysis of covariance; after adjustment, the direction and statistical significance of the between-group differences were unchanged. The combination group showed a larger post-treatment decline in CA125 than the control group (P < 0.001), whereas the between-group difference in SCC-Ag change was not significant (P = 0.866). With respect to immune function, the combination group showed a decrease in CD4^+^ T cells and an increase in CD8^+^ T cells, both significantly different from the control group (P < 0.001).

**Table 1.**
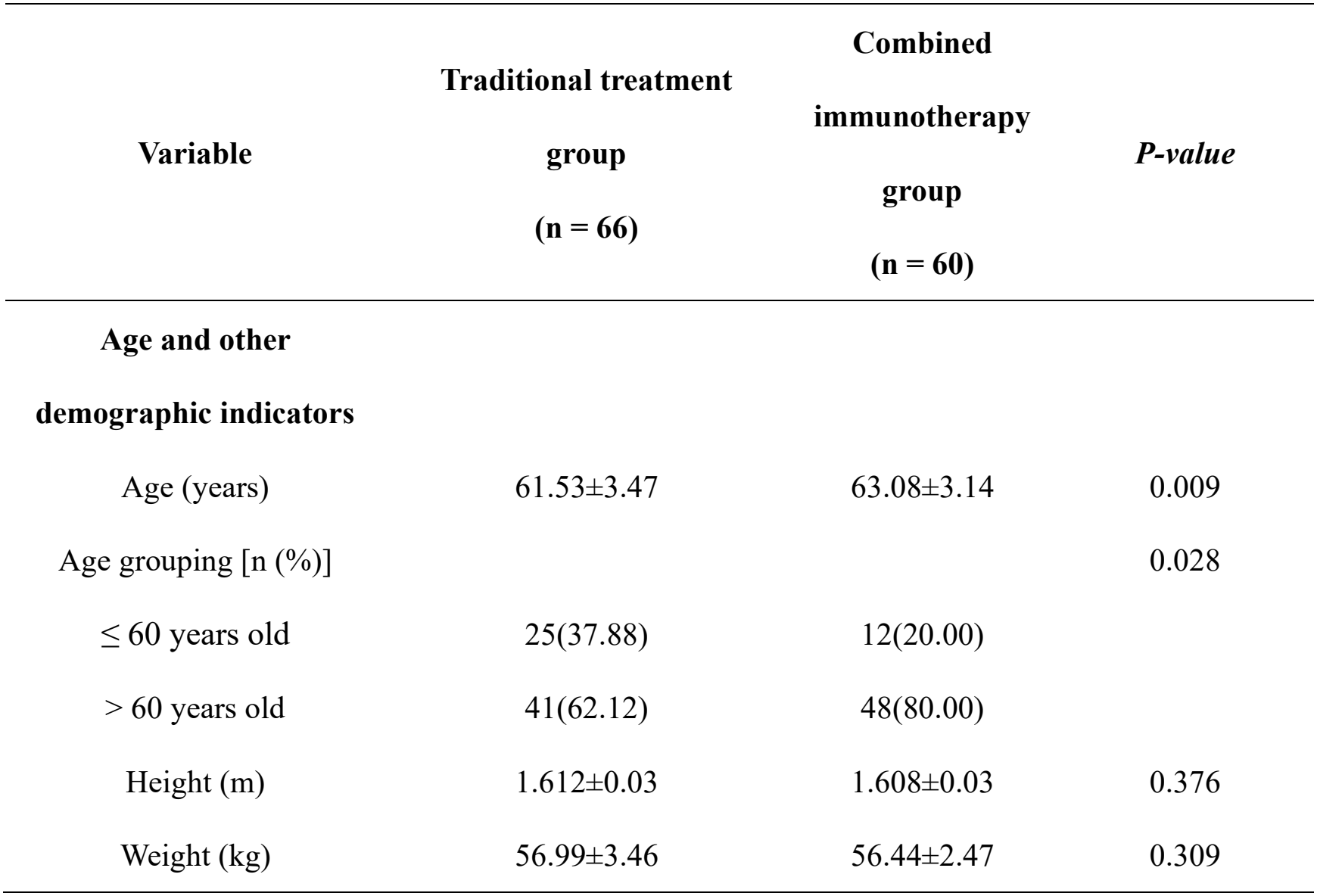
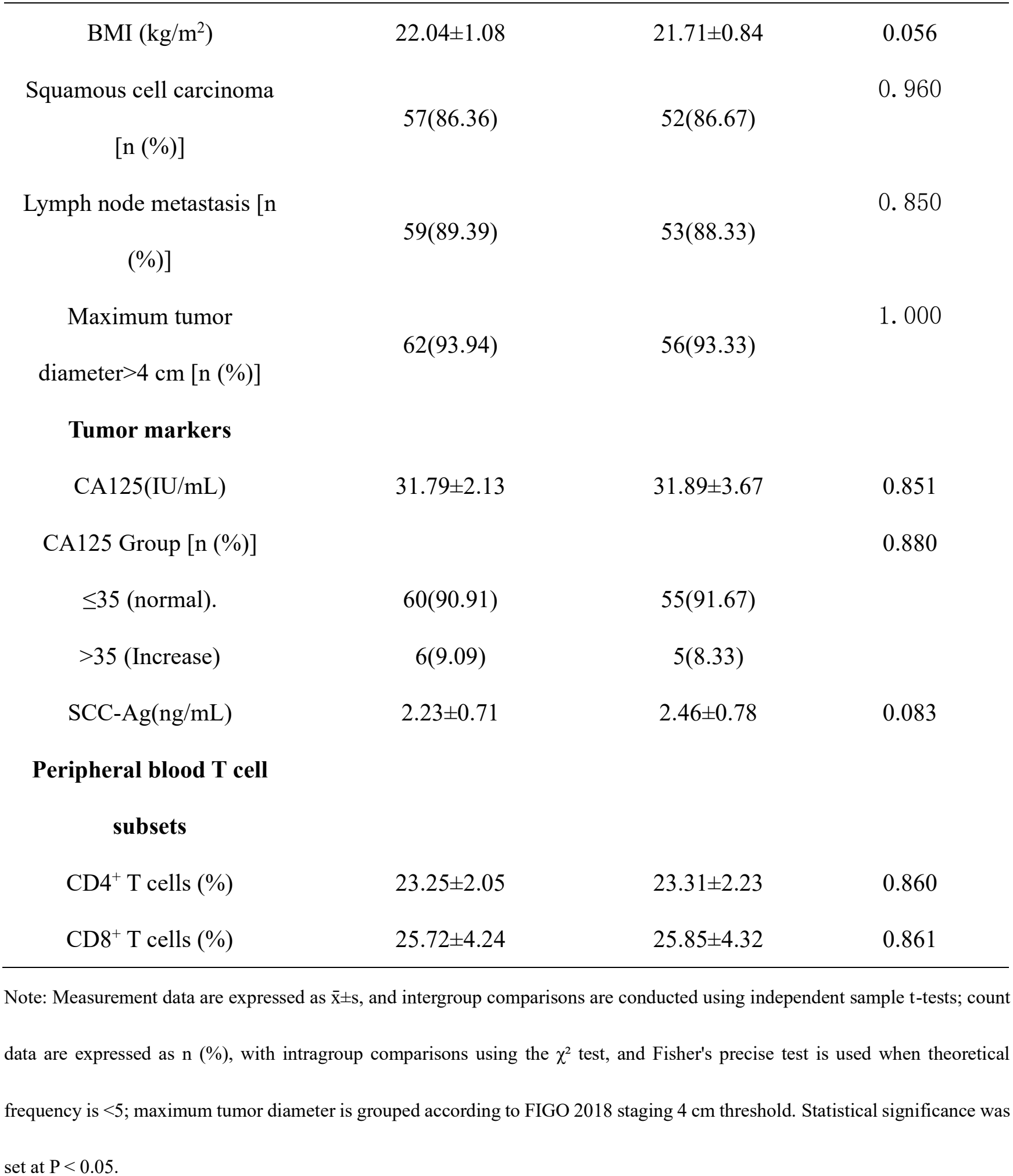
Comparison of baseline characteristics between the two groups.

| **Variable** | **Traditional treatment group**<br>**(n = 66)** | **Combined immunotherapy group**<br>**(n = 60)** | ***P-value*** |
|---|---|---|---|
| **Age and other demographic indicators** |  |  |  |
| Age (years) | 61.53±3.47 | 63.08±3.14 | 0.009 |
| Age grouping [n (%)] |  |  | 0.028 |
| ≤ 60 years old | 25(37.88) | 12(20.00) |  |
| > 60 years old | 41(62.12) | 48(80.00) |  |
| Height (m) | 1.612±0.03 | 1.608±0.03 | 0.376 |
| Weight (kg) | 56.99±3.46 | 56.44±2.47 | 0.309 |
| BMI (kg/m<sup>2</sup>) | 22.04±1.08 | 21.71±0.84 | 0.056 |
| Squamous cell carcinoma [n (%)] | 57(86.36) | 52(86.67) | 0．960 |
| Lymph node metastasis [n (%)] | 59(89.39) | 53(88.33) | 0．850 |
| Maximum tumor diameter>4 cm [n (%)] | 62(93.94) | 56(93.33) | 1．000 |
| **Tumor markers** |  |  |  |
| CA125(IU/mL) | 31.79±2.13 | 31.89±3.67 | 0.851 |
| CA125 Group [n (%)] |  |  | 0.880 |
| ≤35 (normal). | 60(90.91) | 55(91.67) |  |
| >35 (Increase) | 6(9.09) | 5(8.33) |  |
| SCC-Ag(ng/mL) | 2.23±0.71 | 2.46±0.78 | 0.083 |
| **Peripheral blood T cell subsets** |  |  |  |
| CD4<sup>+</sup> T cells (%) | 23.25±2.05 | 23.31±2.23 | 0.860 |
| CD8<sup>+</sup> T cells (%) | 25.72±4.24 | 25.85±4.32 | 0.861 |

Note: Measurement data are expressed as x̄±s, and intergroup comparisons are conducted using independent sample t-tests; count data are expressed as n (%), with intragroup comparisons using the χ² test, and Fisher's precise test is used when theoretical frequency is <5; maximum tumor diameter is grouped according to FIGO 2018 staging 4 cm threshold. Statistical significance was set at P < 0.05.

**Table 2.** Comparison of peripheral hematologic tumor markers and T cell test results before and after treatment in both groups.

|  | Traditional treatment group<br>(n = 66) |  |  | Combined immunotherapy group<br>(n = 60) |  |  |  |
|---|---|---|---|---|---|---|---|
| Parameters | Before treatment | After treatment | d<sub>1</sub> | Before treatment | After treatment | d<sub>2</sub> | Intergroup *P* |
| SCC-Ag (ng/mL) | 2.23 ± 0.71 | 2.19 ± 0.72 | -0.04 ± 0.15 | 2.46 ± 0.78 | 2.41 ± 0.75 | -0.049 ± 0.15 | 0.866 |
| CA125 (IU/mL) | 31.79 ± 2.13 | 31.77 ± 2.21 | -0.02 ± 0.51 | 31.89 ± 3.67 | 31.57 ± 3.68 | -0.32 ± 0.19 | <0.001<sup>*</sup> |
| CD4<sup>+</sup>T cells (%) | 23.25 ± 2.05 | 23.31 ± 2.11 | 0.07 ± 0.35 | 23.31 ± 2.23 | 22.96 ± 2.14 | -0.350 ± 0.448 | <0.001<sup>*</sup> |
| CD8<sup>+</sup>T cells (%) | 25.72 ± 4.24 | 25.70 ± 4.27 | -0.01 ± 0.14 | 25.85 ± 4.32 | 26.18 ± 4.48 | 0.33 ± 0.43 | <0.001<sup>*</sup> |
| Efficacy | Traditional treatment group (n = 66) | Combined immunotherapy group (n = 60) | *P* |  |  |  |  |
| CR | 22(33.33%) | 27(45.00%) | 0.203 |  |  |  |  |
| PR | 21(31.82%) | 22(36.67%) | 0.579 |  |  |  |  |
| SD | 16(24.24%) | 9(15.00%) | 0.264 |  |  |  |  |
| PD | 7(10.61%) | 2(3.33%) | 0.168 |  |  |  |  |
| ORR | 43(65.15%) | 49(81.67%) | 0.045<sup>*</sup> |  |  |  |  |
| DCR | 59(89.39%) | 58(96.67%) | 0.168 |  |  |  |  |

Note: Measurement data are expressed as x̄±s. d<sub>1</sub> and d<sub>2</sub> represent the pre- and post-treatment differences between the traditional and combination immunotherapy groups, respectively, where d = after treatment − before treatment; intragroup comparisons were measured using paired sample t-tests, and between-group comparisons were conducted using independent sample t-tests (Welch correction was used for variance discrepancies); \* represents P<0.05, indicating statistically significant differences.
Note: ∗ represents P<0.05, indicating statistically significant differences; counting data group comparisons were conducted using Fisher's exact test; PD and DCR are complements, and the test P values are the same.

Table 2 Comparison of peripheral hematologic tumor markers and T cell test results before and after treatment in both groups

### 2.3 Comparison of short-term imaging RECIST 1.1 efficacy after treatment between the two groups

Objective tumor response was assessed according to RECIST 1.1, with ORR = CR + PR and DCR = CR + PR + SD. In the traditional concurrent chemoradiotherapy group (n = 66), there were 22 CR, 21 PR, 16 SD, and 7 PD cases; ORR = (22 + 21)/66 = 65.15% and DCR = (22 + 21 + 16)/66 = 89.39%. In the concurrent chemoradiotherapy plus sintilimab group (n = 60), there were 27 CR, 22 PR, 9 SD, and 2 PD cases; ORR = (27 + 22)/60 = 81.67% and DCR = (27 + 22 + 9)/60 = 96.67%. The between-group difference in ORR was statistically significant (P = 0.045), whereas the difference in DCR was not (P = 0.168) (Table 3).

**Table 3.** Comparison of short-term imaging efficacy after treatment between the two groups.

| Efficacy | Traditional treatment group (n = 66) | Combined immunotherapy group (n = 60) | *P* |
|---|---|---|---|
| CR | 22(33.33%) | 27(45.00%) | 0.203 |
| PR | 21(31.82%) | 22(36.67%) | 0.579 |
| SD | 16(24.24%) | 9(15.00%) | 0.264 |
| PD | 7(10.61%) | 2(3.33%) | 0.168 |
| ORR | 43(65.15%) | 49(81.67%) | 0.045<sup>*</sup> |
| DCR | 59(89.39%) | 58(96.67%) | 0.168 |

Note: \* represents P<0.05, indicating statistically significant differences; counting data group comparisons were conducted using Fisher's exact test; PD and DCR are complements, and the test P values are the same.

### 2.4 Survival analysis

Median PFS was 10.0 months in the CCRT group. Adding sintilimab extended this to 18.6 months, an absolute difference of 8.6 months, and the separation between the two Kaplan–Meier curves was confirmed by the log-rank test (χ^2^ = 13.015, P = 0.0003). Overall survival showed a similar pattern: median OS was 27.3 months in the CCRT group and was not reached in the CCRT plus sintilimab group during the observation period, corresponding to a statistically significant survival advantage (χ^2^ = 9.005, P = 0.0027) (Figure 2).

**Figure 2.**
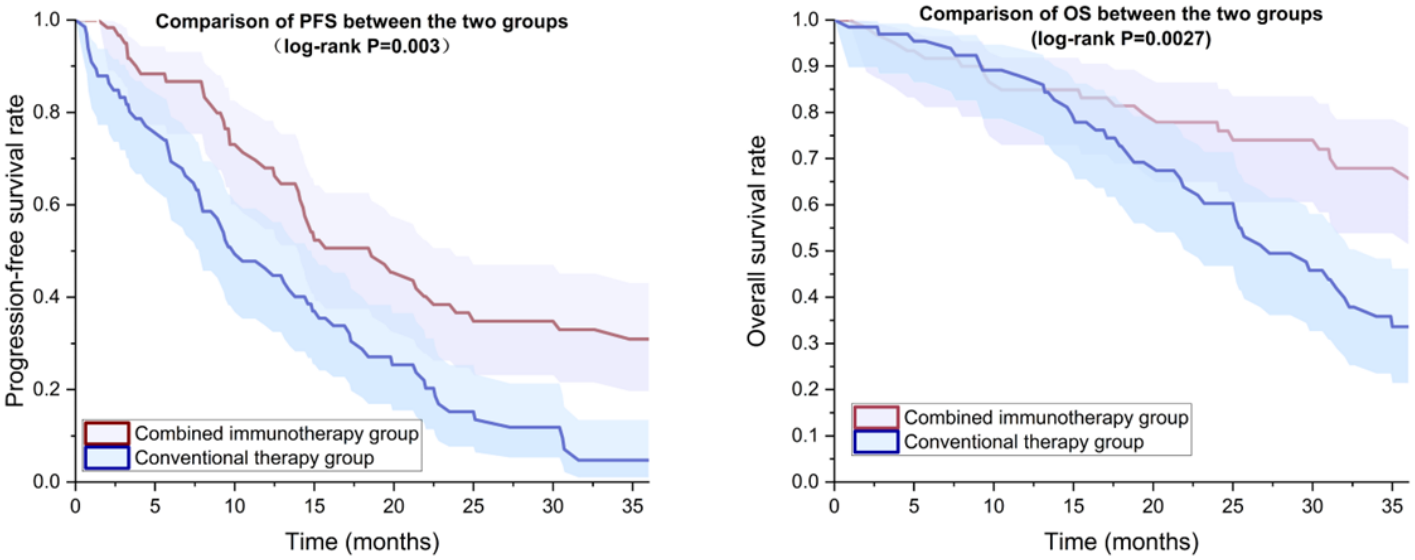
Kaplan-Meier survival curves for PFS and OS in both groups

Using the median post-treatment CA125 level (31.25 IU/mL), we divided the 126 patients into low-level (62 cases) and high-level (64 cases) groups. Median PFS was 19.4 months in the low-level group and 11.9 months in the high-level group (HR = 1.89, 95% CI: 1.26–2.83; log-rank χ^2^ = 9.880, P = 0.0017), and the 24-month PFS rates were 39.4% and 12.0%, respectively (Figure 3A). OS showed a comparable pattern: median OS was not reached in the low-level group, compared with 31.5 months in the high-level group, but this difference was not statistically significant (HR 1.63, 95% CI 0.96–2.77, P = 0.070; Figure 3C).

**Figure 3.**
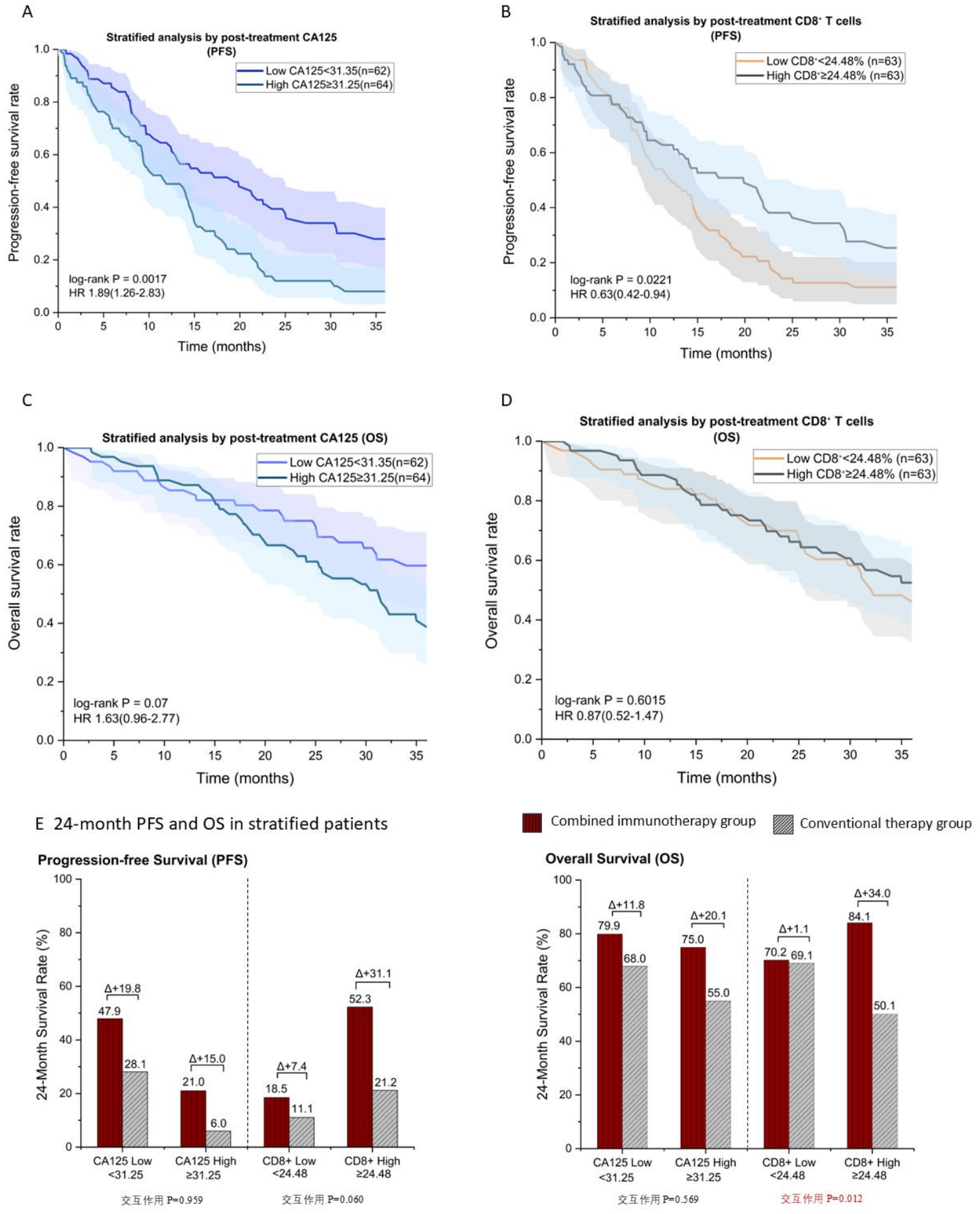
Survival analysis of CA125 and CD8^+^T cell stratification after treatment

Patients were divided into high and low groups by the median post-treatment CD8^+^ T-cell proportion (24.48%), with 63 patients in each. Median PFS was 19.9 months in the high-proportion group versus 12.4 months in the low-proportion group (HR 0.63, 95% CI 0.42–0.94; log-rank χ^2^ = 5.238, P = 0.0221; Figure 3B). For OS, the curves were similar, with 12-, 24-, and 30-month rates of 88.6% vs 84.0%, 68.0% vs 69.9%, and 60.7% vs 58.3%, respectively (no significant difference; HR 0.87, 95% CI 0.52–1.47; log-rank χ^2^ = 0.273, P = 0.602). Median OS was not reached in the high-proportion group, whereas it was 32.3 months in the low-proportion group (Figure 3D). Taken together, these data suggest that a higher post-treatment CD8^+^ T-cell proportion is associated with better PFS, marking it as a potential prognostic indicator.

After stratification by the median post-treatment CD8^+^ T-cell proportion (24.48%) and CA125 level (31.25 IU/mL), we compared the 24-month survival rates of the two treatments within each stratum. In the high-CD8^+^ stratum, the combination arm reported 24-month PFS and OS rates of 52.3% and 84.1%, respectively, both higher than those of the CCRT control arm (21.2% and 50.1%). The two arms were more closely matched in the low-proportion stratum: 24-month PFS was 18.5% versus 11.1%, and OS 70.2% versus 69.1%. CD8^+^ T-cell stratification interacted significantly with treatment arm for OS (P = 0.012), with a borderline result for PFS (P = 0.060). By contrast, CA125 showed no treatment-by-subgroup interaction in either OS (P = 0.569) or PFS (P = 0.959) (Figure 3E).

To test the robustness of these findings, we performed a 3-month landmark analysis (n = 112). In this subset, CD8^+^ T-cell stratification significantly improved PFS discrimination (P = 0.0024). After excluding early events, the PFS and OS benefits of combination immunotherapy remained significant (HR = 0.53 and 0.47, respectively). RMST analysis further showed a longer mean PFS in the high-CD8^+^ group than in the low-CD8^+^ group (24.4 vs. 16.0 months), whereas high CA125 was associated with a shorter PFS (17.8 vs. 22.5 months). These results were consistent with the primary analysis, supporting the robustness of our conclusions against early events and deviations from the proportional hazards assumption.

We used a Cox proportional hazards model to assess predictors of PFS. In addition to age and BMI, we included treatment group, post-treatment CA125, and CD8^+^ T-cell counts as covariates, with continuous variables scaled per one-standard-deviation increase. The univariate and multivariable hazard ratios are summarized in Table 4.

**Table 4.** Univariate and multivariate Cox regression analyses of progression-free survival and overall survival in both groups.

| Variable | One-factor HR (95% CI) | *P* value | Multifactor HR (95% CI) | *P* value |
|---|---|---|---|---|
| **PFS events: 99 cases / n=126** |  |  |  |  |
| Treatment Groups (Combined Immunotherapy vs. Traditional Treatment) | 0.48 (0.32−0.72) | <0.001<sup>*</sup> | 0.44 (0.29−0.68) | <0.001<sup>*</sup> |
| Age (per 1 SD = 3.39 years) | 0.93 (0.75−1.14) | 0.480 | 0.97 (0.78−1.20) | 0.755 |
| BMI(per 1 SD = 0.98 kg/m²) | 0.93 (0.76−1.13) | 0.458 | 0.85 (0.69−1.04) | 0.109 |
| After treatment, CA125 (per 1 SD =2.99 IU/mL) | 1.11 (0.97−1.28) | 0.121 | 1.13 (0.96−1.32) | 0.136 |
| After treatment, CD8<sup>+</sup>T cells were used (per 1 SD = 4.36%) | 0.87 (0.70−1.08) | 0.214 | 0.93 (0.75−1.15) | 0.506 |
| **OS events: 57 cases / n=126** |  |  |  |  |
| Treatment Groups (Combined Immunotherapy vs. Traditional Treatment) | 0.44 (0.25−0.76) | 0.004<sup>*</sup> | 0.41 (0.23−0.73) | 0.002<sup>*</sup> |
| Age (per 1 SD = 3.39 years) | 1.07 (0.81−1.40) | 0.638 | 1.16 (0.87−1.54) | 0.317 |
| BMI (per 1 SD = 0.98 kg/m²) | 1.03 (0.80−1.32) | 0.840 | 0.96 (0.74−1.24) | 0.736 |
| After treatment, CA125(per 1 SD =2.99 IU/mL) | 1.09 (0.89−1.33) | 0.421 | 1.07 (0.83−1.38) | 0.593 |
| After treatment, CD8<sup>+</sup>T cells were used (per 1 SD = 4.36%) | 0.95 (0.73−1.24) | 0.716 | 0.99 (0.76−1.29) | 0.949 |

Note: HR, hazard ratio; CI, confidence interval; PFS, progression-free survival; OS, overall survival; SD, standard deviation; Traditional treatment = traditional concurrent chemoradiotherapy (control group); After treatment, CA125 was included as a continuous variable per 1 SD; In sensitivity analysis, when included by median binary classification, HR=1.66 (95% CI: 1.09~2.53, P = 0.018), with both parameterization directions consistent.

Treatment group was the only independent influencing factor for PFS: the univariate HR was 0.48 (95% CI: 0.32–0.72, P < 0.001) and the multivariable HR 0.44 (95% CI: 0.29–0.68, P < 0.001). The estimates were similar in the two models, and combination immunotherapy was associated with a 56% lower hazard of disease progression or death. Age (HR = 0.97, 95% CI: 0.78–1.20, P = 0.755), BMI (HR = 0.85, 95% CI: 0.69–1.04, P = 0.109), post-treatment CA125 (HR = 1.13, 95% CI: 0.96–1.32, P = 0.136), and post-treatment CD8^+^ T cells (HR = 0.93, 95% CI: 0.75–1.15, P = 0.506) were not independent prognostic factors for PFS. The same Cox model was then fitted with OS as the outcome. Treatment group remained an independent prognostic factor for OS: the univariate HR was 0.44 (95% CI 0.25–0.76, P = 0.004) and the multivariable HR 0.41 (95% CI 0.23–0.73, P = 0.002), corresponding to a 59% lower mortality hazard. Age (HR = 1.16, P = 0.317), BMI (HR = 0.96, P = 0.736), post-treatment CA125 (HR = 1.07, P = 0.593), and post-treatment CD8^+^ T cells (HR = 0.99, P = 0.949) showed no independent association with OS. The C-indices for the two models were 0.618 and 0.611, indicating moderate discrimination. The Schoenfeld residual test showed no violation of the proportional hazards assumption for any variable (P > 0.05 for each).

After adjustment, treatment group was the only independent predictor of both PFS and OS. Age, BMI, and CA125 level were not significant predictors after adjustment. CD8^+^ T-cell counts showed a trend toward significance in the unadjusted PFS model but lost their signal after adjustment. Rather than acting as standalone predictors, CD8^+^ T cells appeared to function as treatment effect modifiers: they significantly influenced the OS benefit from combination therapy (P = 0.012), with a similar trend for PFS (P = 0.060). CA125 stratification showed no such interaction.

Two additional analyses were performed to test the robustness of the findings. Excluding early progressions in the 3-month landmark analysis did not alter the survival advantage of CCRT plus sintilimab. Similarly, RMST at 36 months confirmed a consistent benefit irrespective of the hazards assumption. Together, these analyses indicate that the main finding is robust and not driven by early events.

## 3. Discussion

In this retrospective cohort, we compared CCRT alone with CCRT plus sintilimab in patients with locally advanced cervical cancer. The combination regimen produced a higher objective response rate and meaningful improvements in both PFS and OS, gains that persisted after adjustment for age, BMI, and post-treatment biomarker levels in the multivariable Cox model. These hazard ratios, less than half those with CCRT alone, indicate that the magnitude of benefit was not merely a reflection of baseline differences between the groups.

A particularly informative finding concerned the immune microenvironment. Patients whose post-treatment CD8^+^ T-cell proportion exceeded the median had notably longer PFS, and interaction analysis showed that CD8^+^ T-cell stratification specifically modified the treatment effect on OS (P = 0.012). This pattern suggests a biological gradient: patients with a more robust CD8^+^ T-cell response after CCRT appeared to derive the greatest survival advantage from PD-1 blockade. Post-treatment CA125, by contrast, stratified PFS as expected, with elevated levels predicting shorter survival, but showed no interaction with treatment assignment, suggesting that CA125 reflects overall tumor burden rather than treatment-specific responsiveness.

Taken together, these results indicate that adding a PD-1 inhibitor to CCRT is associated with improved tumor control and survival in this population. The post-treatment CD8^+^ T-cell proportion emerges as a promising candidate for efficacy stratification that warrants prospective validation. Given the retrospective design and limited sample size, however, we caution against adopting it as a standalone criterion for treatment decisions at this stage.

Concurrent chemoradiotherapy remains the backbone of radical management for LACC^19^. The ORR, PFS, and OS advantages observed in our combination arm are consistent with those reported in KEYNOTE-A18, a randomized double-blind phase III trial. In high-risk LACC, pembrolizumab given with CCRT followed by continued maintenance improved PFS, and later analyses of that trial also demonstrated an OS benefit^20,21^. Consistent with these findings, Sukhin et al.^22^ similarly observed improvements in both progression-free survival (PFS) and overall survival (OS) when immune checkpoint inhibitors were combined with concurrent chemoradiotherapy. This consistency supports the clinical hypothesis that PD-1 pathway inhibition on a background of chemoradiotherapy may enhance disease control. The two studies differed in their intervention backgrounds: KEYNOTE-A18 administered treatment continuously during both the concurrent and maintenance phases. In our retrospective single-center cohort, sintilimab was also given continuously, including throughout the maintenance phase. Accordingly, the observed survival differences should be regarded as observational associations within this treatment setting rather than causal evidence. Causal conclusions, as well as direct comparisons between different PD-1 inhibitors, remain to be confirmed in randomized controlled trials.

Radiotherapy and immunotherapy can act synergistically against tumors. Radiation kills cancer cells and releases antigens that help activate effector T cells, but it also triggers immunosuppressive feedback such as PD-L1 upregulation^23^. PD-1 blockade then partially rescues and sustains the effector function of tumor-specific T cells^24–26^. Studies of cervical squamous cell carcinoma specimens have linked post-radiotherapy intratumoral CD8^+^ T-cell infiltration and altered PD-L1 expression to clinical outcomes^27^. In our cohort, patients receiving combination therapy showed a clear post-treatment immune shift, with CD8^+^ T-cell proportions rising and CD4^+^ proportions falling, consistent with immune remodeling. We also found a significant interaction between CD8^+^ stratification and treatment for OS (P = 0.012), indicating that patients who mount a stronger CD8^+^ response are likely to benefit most from sintilimab. Peripheral blood, however, captures only part of the picture: it cannot reflect intratumoral features such as T-cell density, clonality, or exhaustion, and it is readily influenced by treatment-related lymphopenia and sampling timing^28,29^. Thus, although we observed an elevated CD8^+^ proportion, whether this reflects an intratumoral shift that enhances PD-1 blockade still requires confirmation through direct analysis of tumor-infiltrating lymphocytes.

The interaction between post-treatment CD8^+^ T-cell stratification and OS provides preliminary evidence for its role as a candidate effector modifier. Available evidence indicates that the response to PD-1 blockade is governed not simply by the abundance of individual immune cell populations but by the equilibrium between effector and immunosuppressive T-cell subsets^30^. In cervical cancer, the tumor immune microenvironment and post-treatment immune status also show substantial heterogeneity^31^. A higher proportion of post-treatment CD8^+^ T cells both reflects a favorable systemic immune status and serves as a correlate of treatment response. When CD8^+^ T cells were dichotomized at the median, they did not emerge as independent prognostic factors for either PFS or OS in the multivariable Cox model, and for PFS the interaction with treatment arm reached only borderline significance (P = 0.060). Accordingly, the post-treatment CD8^+^ T-cell proportion is currently best regarded as an exploratory stratification indicator; confirmation will require validation in independent prospective cohorts using preset cutoffs, together with tumor-tissue CD8^+^ T cells, PD-L1, HPV-related markers, and T-cell functional phenotyping.

In this study, higher post-treatment CA125 levels were associated with shorter PFS, but there was no interaction with treatment modality, suggesting that CA125 reflects tumor burden, treatment response, or overall prognosis rather than specifically predicting benefit from PD-1 inhibitors. In selected histological subtypes such as cervical adenocarcinoma, CA125 has been associated with unfavorable pathological characteristics and elevated recurrence risk^32^; however, its prognostic utility in the broader LACC population receiving definitive CCRT remains inadequately established. Here, the absolute change in CA125 was small, and CA125 can be influenced by non-neoplastic gynecological or peritoneal factors, limiting its value as an independent basis for selecting patients for immunotherapy.

Clinically, this study adds real-world response and survival data for PD-1 inhibition combined with CCRT in locally advanced cervical cancer and introduces longitudinal peripheral immune markers to help explain why patients respond differently. Conceptually, our results support a working model in which radiation releases tumor antigens, these provoke a T-cell response, and PD-1 blockade releases that response from inhibitory restraint, offering a tractable framework for individualized stratification. In summary, adding sintilimab to CCRT improved the objective response rate and prolonged both PFS and OS. Among the peripheral markers we tracked, the post-treatment CD8^+^ T-cell proportion was most closely linked to treatment benefit, whereas CA125 reflected tumor burden without distinguishing who would gain from immunotherapy. Neither marker should be used alone to guide treatment decisions. Several limitations warrant consideration. As a single-center retrospective study without randomization, it cannot exclude treatment selection bias or residual confounding. Although we adjusted for age and BMI, the age distribution remained uneven between groups, and key variables such as FIGO stage, tumor volume, nodal involvement, and comorbidities were not included in the models. The modest sample size also limited the statistical power of subgroup analyses, and the cutoffs for CD8^+^ and CA125 were derived from the data rather than set in advance. In addition, because immune markers were sampled only before and after treatment, we could not capture the dynamics in between or assess T-cell exhaustion. Adverse events were not formally compared between groups, which limits our safety conclusions.

Future studies should confirm these findings in multicenter prospective cohorts with randomized or matched controls. Equally important is the integration of longitudinal blood sampling with tumor-tissue profiling to determine whether peripheral CD8^+^ changes truly mirror events within the tumor. Only then can these biomarkers be validated for clinical use.

## Data Availability

The data that support the findings of this study are available from the corresponding author upon reasonable request.

## Contributors

Conceptualization: Yanjun Xue; Investigation: Yanjun Xue; Data curation: Yingqi Hao, Chaoli Shi, Xiaohui Sun, Kaili Li, Jinmeng Li; Formal analysis: Yingqi Hao; Writing-original draft: Yingqi Hao; Writing – review & editing: Yanjun Xue; Supervision: Yanjun Xue. All authors read and approved the final manuscript.

## References

1. Behtash, N., Izadimood, N., Teymoordash, S. N. & Ramhormozian, S. Global incidence of cervical cancer. in Comprehensive Review of Lower Genital Neoplasms 5–8 (Elsevier, 2026). doi:10.1016/B978-0-443-44564-4.00010-3.

2. Bhatla, N. et al. Revised FIGO staging for carcinoma of the cervix uteri. Intl J Gynecology & Obste 145, 129–135 (2019).

3. Kornovski, I., Gorchev, G. & Iakov, M. [Therapeutic approach in locally advanced cervical cancer]. Akush Ginekol (Sofiia) 45, 20–24 (2006).

4. Rose, P. G. et al. Concurrent Cisplatin-Based Radiotherapy and Chemotherapy for Locally Advanced Cervical Cancer. N Engl J Med 340, 1144–1153 (1999).

5. Keys, H. M. et al. Cisplatin, Radiation, and Adjuvant Hysterectomy Compared with Radiation and Adjuvant Hysterectomy for Bulky Stage IB Cervical Carcinoma. N Engl J Med 340, 1154–1161 (1999).

6. Morris, M. et al. Pelvic Radiation with Concurrent Chemotherapy Compared with Pelvic and Para-Aortic Radiation for High-Risk Cervical Cancer. N Engl J Med 340, 1137–1143 (1999).

7. Whitney, C. W. et al. Randomized Comparison of Fluorouracil Plus Cisplatin Versus Hydroxyurea as an Adjunct to Radiation Therapy in Stage IIB-IVA Carcinoma of the Cervix With Negative Para-Aortic Lymph Nodes: A Gynecologic Oncology Group and Southwest Oncology Group Study. JCO 17, 1339–1339 (1999).

8. Ye, J. et al. Assessing the Magnitude of Immunogenic Cell Death Following Chemotherapy and Irradiation Reveals a New Strategy to Treat Pancreatic Cancer. Cancer Immunol Res 8, 94–107 (2020).

9. Suzuki, Y. et al. Immunogenic Tumor Cell Death Induced by Chemoradiotherapy in Patients with Esophageal Squamous Cell Carcinoma. Cancer Research 72, 3967–3976 (2012).

10. Ribas, A. & Wolchok, J. D. Cancer immunotherapy using checkpoint blockade. Science 359, 1350–1355 (2018).

11. Sharabi, A. B., Lim, M., DeWeese, T. L. & Drake, C. G. Radiation and checkpoint blockade immunotherapy: radiosensitisation and potential mechanisms of synergy. The Lancet Oncology 16, e498–e509 (2015).

12. Colombo, N. et al. Pembrolizumab for Persistent, Recurrent, or Metastatic Cervical Cancer. N Engl J Med 385, 1856–1867 (2021).

13. Oaknin, A. et al. Atezolizumab plus bevacizumab and chemotherapy for metastatic, persistent, or recurrent cervical cancer (BEATcc): a randomised, open-label, phase 3 trial. The Lancet 403, 31–43 (2024).

14. Knoth, J. et al. Distant Metastasis After Chemoradiation and Image Guided Adaptive Brachytherapy in Locally Advanced Cervical Cancer. International Journal of Radiation Oncology*Biology*Physics 123, 503–512 (2025).

15. Hiam-Galvez, K. J., Allen, B. M. & Spitzer, M. H. Systemic immunity in cancer. Nat Rev Cancer 21, 345–359 (2021).

16. Wu, Y. et al. Clinical significance of peripheral blood and tumor tissue lymphocyte subsets in cervical cancer patients. BMC Cancer 20, 173 (2020).

17. Von Elm, E. et al. The Strengthening the Reporting of Observational Studies in Epidemiology (STROBE) Statement: Guidelines for Reporting Observational Studies. PLoS Med 4, e296 (2007).

18. Eisenhauer, E. A. et al. New response evaluation criteria in solid tumours: Revised RECIST guideline (version 1.1). European Journal of Cancer 45, 228–247 (2009).

19. Cibula, D. et al. ESGO/ESTRO/ESP Guidelines for the management of patients with cervical cancer – Update 2023. Radiotherapy and Oncology 184, 109682 (2023).

20. Lorusso, D. et al. Pembrolizumab or placebo with chemoradiotherapy followed by pembrolizumab or placebo for newly diagnosed, high-risk, locally advanced cervical cancer (ENGOT-cx11/GOG-3047/KEYNOTE-A18): a randomised, double-blind, phase 3 clinical trial. The Lancet 403, 1341–1350 (2024).

21. Lorusso, D. et al. Pembrolizumab or placebo with chemoradiotherapy followed by pembrolizumab or placebo for newly diagnosed, high-risk, locally advanced cervical cancer (ENGOT-cx11/GOG-3047/KEYNOTE-A18): overall survival results from a randomised, double-blind, placebo-controlled, phase 3 trial. The Lancet 404, 1321–1332 (2024).

22. Sukhin, V. S. et al. 1170P Pembrolizumab (Pembro) + concurrent chemoradiotherapy (CCRT) for high-risk locally advanced cervical cancer (LACC): Post hoc subgroup analyses from the phase III ENGOT-cx11/GOG-3047/KEYNOTE-A18 study. Annals of Oncology 36, S723 (2025).

23. Deng, L. et al. Irradiation and anti–PD-L1 treatment synergistically promote antitumor immunity in mice. J. Clin. Invest. 124, 687–695 (2014).

24. Barber, D. L. et al. Restoring function in exhausted CD8 T cells during chronic viral infection. Nature 439, 682–687 (2006).

25. Im, S. J. et al. Defining CD8+ T cells that provide the proliferative burst after PD-1 therapy. Nature 537, 417–421 (2016).

26. Tumeh, P. C. et al. PD-1 blockade induces responses by inhibiting adaptive immune resistance. Nature 515, 568–571 (2014).

27. Mori, Y. et al. Analysis of radiotherapy-induced alteration of CD8+ T cells and PD-L1 expression in patients with uterine cervical squamous cell carcinoma. Oncol Lett 21, 446 (2021).

28. Holub, K., Vargas, A. & Biete, A. Radiation-induced lymphopenia: the main aspects to consider in immunotherapy trials for endometrial and cervical cancer patients. Clin Transl Oncol 22, 2040–2048 (2020).

29. Cho, O. et al. Prognostic implication of simultaneous anemia and lymphopenia during concurrent chemoradiotherapy in cervical squamous cell carcinoma. Tumour Biol 39, 1010428317733144 (2017).

30. Kumagai, S. et al. The PD-1 expression balance between effector and regulatory T cells predicts the clinical efficacy of PD-1 blockade therapies. Nat Immunol 21, 1346–1358 (2020).

31. Zhang, Y. et al. Baseline immunity and impact of chemotherapy on immune microenvironment in cervical cancer. Br J Cancer 124, 414–424 (2021).

32. Kim, N. et al. Significance of serum CA125 level in surgically resected cervical adenocarcinoma with adverse features. J Gynecol Oncol 32, e72 (2021).

